# Role of the complement system in Long COVID

**DOI:** 10.1101/2024.03.14.24304224

**Authors:** Vadim Farztdinov, Boris Zühlke, Franziska Sotzny, Fridolin Steinbeis, Martina Seifert, Claudia Kedor, Kirsten Wittke, Pinkus Tober-Lau, Thomas Zoller, Kathrin Textoris-Taube, Daniela Ludwig, Clemens Dierks, Dominik Bierbaum, Leif Erik Sander, Leif G Hanitsch, Martin Witzenrath, Florian Kurth, Michael Mülleder, Carmen Scheibenbogen, Markus Ralser

**Affiliations:** Core Facility High Throughput Mass Spectrometry, Charité - Universitätsmedizin Berlin, corporate member of Freie Universität Berlin and Humboldt-Universität zu Berlin, Berlin, Germany; Institute of Biochemistry, Charité - Universitätsmedizin Berlin, corporate member of Freie Universität Berlin and Humboldt-Universität zu Berlin, Berlin, Germany; Institute of Medical Immunology, Charité - Universitätsmedizin Berlin, corporate member of Freie Universität Berlin and Humboldt Universität zu Berlin, Berlin, Germany; Department of Infectious Diseases, Respiratory Medicine and Critical Care, Charité - Universitätsmedizin Berlin, corporate member of Freie Universität Berlin and Humboldt-Universität zu Berlin, Berlin, Germany; German Center for Lung Research (DZL), Berlin, Germany; Centre for Human Genetics, Nuffield Department of Medicine, University of Oxford, UK; Max Planck Institute for Molecular Genetics, Berlin Germany

**Author notes:** These authors contributed equally to this work (Vadim Farztdinov, Boris Zühlke).

## Abstract

Long COVID, or Post-Acute COVID Syndrome (PACS), may develop following SARS-CoV-2 infection, posing a substantial burden to society. Recently, PACS has been linked to a persistent activation of the complement system (CS), offering hope for both a diagnostic tool and targeted therapy. However, our findings indicate that, after adjusting proteomics data for age, body mass index and sex imbalances, the evidence of complement system activation disappears. Furthermore, proteomic analysis of two orthogonal cohorts—one addressing PACS following severe acute phase and another after a mild acute phase—fails to support the notion of persistent CS activation. Instead, we identify a proteomic signature indicative of either ongoing infections or sustained immune activation similar to that observed in acute COVID-19, particularly within the mild-PACS cohort.

## Main

Long COVID or Post acute COVID Syndrome (PACS) may emerge upon SARS-CoV-2 infection, causing a considerable societal burden (*1–3*). The underlying mechanisms are complex with evidence for immune activation and dysregulation, autoantibodies, vascular perfusion and mitochondrial disturbance and viral persistence or reactivation playing a role (*4*). There is no causal therapy available yet. Recently, a study by Cervia-Hasler et al. (*5*) thus generated great interest as it linked the disease to a persistent activation of the complement system (CS), and therefore of the innate immune system, providing a causal explanation for PACS. The results raised hopes for a diagnostic tool and a targeted therapy.

Our curiosity was stirred by a result shown in Figure 8b, demonstrating that in their cohort, age and body mass index (BMI) alone predicted PACS with an area under the Receiver Operating Characteristic (ROC) curve of almost 0.8. This result stands in contrast with other studies in the field. While these agree that age and BMI can be risk factors of PACS, age and BMI alone are deemed insufficient to predict which individuals would develop PACS (*6*).

In seeking to explain why age and BMI are such strong predictors of PACS in this study, we noticed that the cohort was substantially imbalanced for age and BMI. The non-PACS control group predominantly consisted of young individuals with a median age of 36 years and a mean BMI of 25, while individuals in the PACS group were considerably older (median age of 58 years) and had a mean BMI of 28.

We were wondering whether this imbalance might have also affected other results. When comparing the mass-spectrometry based proteomes of the control (non-PACS) and the PACS group without balancing for these demographic factors, CS components indeed appear to be up-regulated in PACS (Figure 1A). However, the CS is known to be age and BMI dependent (*7, 8*). In their data analysis, Cervia-Hasler et al. attempt a correction for demographic effects using linear regression (*5*). Nonetheless, age, sex and BMI can have complex interactions with the plasma proteome, and a simple linear model might hence not be sufficient to capture these interactions. To mitigate the age and BMI imbalances, we used a balanced factorial design strategy by splitting all patients in disease groups by age and sex (Figure 1B). Because the cohort was too small to balance all parameters, we excluded those individuals with vastly different age and/or BMI (Supplementary Table 1). While this strategy reduced the cohort, it resulted in a reasonable balancing by age and BMI in subgroups. The final set consisted of 85 individuals (56 controls and + 29 PACS), with approximately the same ratio of PACS to non-PACS as the initial data set. Comparing the proteomes between PACS and non-PACS in this balanced cohort, none of the complement components was significantly changed in PACS, using the same significance level set by Cervia-Hasler et al. (*5*) (Figure 1C). In contrast, this analysis revealed a different set of plasma proteins that emerged as discriminators of PACS. Apart from immunoglobulins, BCHE was significantly higher in males above 48 years of age, Paraoxonase 3 (PON3) was increased in PACS patients and Syntax Binding Protein 5 (STXBP5) was decreased in PACS patients (*9)*.

**Figure 1.**
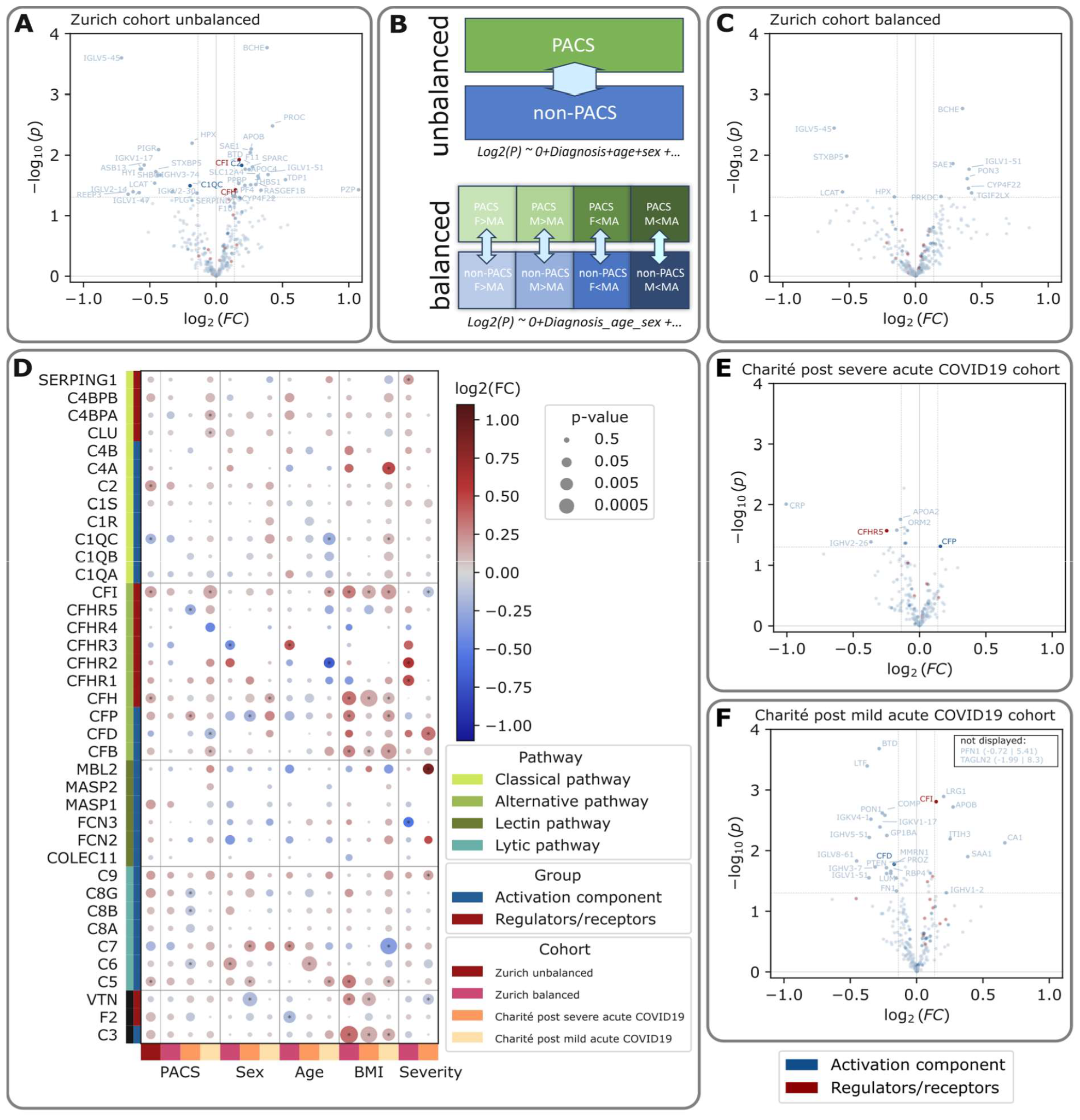
The regulation of the plasma proteome in post severe and mild acute COVID19: Statistical analysis of mass spectrometry proteomics data on the Zurich cohort as described by Cervia-Hasler et al. for the contrast PACS vs. non-PACS, without further balancing for demographic factors. B: Scheme of the factorial design and linear models applied to the Zurich cohort using the linear model log2(P) ∼ 0 + Diagnosis + Age + Sex + Severity versus balanced Zurich, and Charité post severe acute COVID19 and Charité post mild acute COVID19 cohorts using the linear model log(P) ∼ 0 + Diagnosis_age_sex + BMI + Severity considering contrasts between age and sex subgroups, where F=female, m=male, MA=mean age. C) Volcano plot for the contrast PACS vs. non-PACS on the Zurich cohort after balancing and modelling over age and sex subgroups. D) Differential abundance between patients with PACS and recovered patients on the complement system (rows, coloured by pathway and group of the complement system) for the three cohorts (first four columns) and effects of sex, age, BMI and severity during the acute phase; circle sizes represent p-values with an asterisk in the middle if the p-value is below the significance threshold of 0.05, the colour intensity as scaled by the colour bar represents over-/underexpression and log2 fold change of the gene product between the given contrasts. In all volcano plots complement system activation components are coloured in blue, complement system regulators or receptors are coloured in red, other proteins are coloured in grey. Proteins identified to be differentially abundant were labelled. E, F): Volcano plots for the Charité post-severe and Charité post-mild acute COVID syndrome cohort, respectively, following the balanced factorial design contrasting PACS vs. non-PACS. Thresholds for differential abundance of proteins were set at 1.1/-1.1 for the fold change and 0.05 for the p-value. Statistical analysis was performed with limma without adjustment for multiple testing. Genes were assigned to the complement system as activators or regulators/receptors using the HUGO gene nomenclature committee (https://www.genenames.org/), pathways were assigned using the Gene Ontology Term Biological Process (24, 25).

### Abundance of the complement system in independent PACS cohorts

To add confidence to the results, we set out to orthogonally explore for a persistent complement activation in PACS. We chose two independent cohorts, addressing two different patient groups suffering from the syndrome. First, we chose a study cohort that closely resembles the study design by Cervia-Hasler et al. (*5*) and which focuses on individuals that report PACS symptoms at a 6 months follow up, after being discharged from Charité where they have been treated for a severe acute phase (WHO severity grade 3-7) (Charité post severe acute COVID-19 cohort (*10*)). At the 6-month follow-up visit, plasma samples were collected from 130 individuals, of whom 67 reported symptoms consistent with PACS. This cohort was largely balanced for age, sex, and BMI. Importantly, among these individuals, there was no significant correlation between reporting PACS symptoms and age, sex, BMI, or severity in the acute phase (Methods), reflecting other studies investigating PACS (*6*). Using a high-throughput mass spectrometry platform (*11, 12*), we investigated the plasma proteome at the 6 months follow-up. Samples were analysed in a 3-minute water to acetonitrile active gradient on an Agilent Infinity II Sciex TripleTOF 6600 mass spectrometer system operating in ScanningSWATH mode (*13*). Comparing the PACS to the recovered group, we observed either no changes in several detected CS components, or a slight downregulation, at low effect sizes (Figure 1D). The statistical analysis of this cohort revealed among others the C-reactive protein (CRP), Apolipoprotein A-II (APOA2), Alpha-1-acid glycoprotein 2 (ORM2) and the CS regulator Complement factor H-related protein 5 (CFHR5) to be less abundant in PACS (Figure 1E). Only Properdin (CFP), a regulator of the alternative complement pathway, was significantly, albeit only slightly, upregulated in PACS patients. Thus, in another PACS cohort resembling the study (*5*), we can neither confirm that age or BMI are strong predictors of the development of PACS, nor do we confirm a persistent dysregulation of a broad set of complement factors in these subjects.

A great societal burden of PACS comes from individuals which suffered not from a severe, but rather a mild or moderate acute phase (*10, 14*). PACS in these individuals is characterised by a complex phenotype including fatigue, exertional intolerance, post exertional malaise (PEM) and brain fog as key symptoms with a subset having developed the most debilitating form of PACS, Myalgic Encephalomyelitis/Chronic Fatigue Syndrome (ME/CFS) (*15*). These patients are younger and predominantly female. We hence chose a third cohort which focuses on PACS with fatigue and exertional intolerance (“Charité post mild acute COVID19” cohort). This cohort included 53 PACS patients with persisting moderate to severe fatigue with 25 of them fulfilling CCC diagnostic criteria for ME/CFS (*16*). A total of 27 non-PACS, Post Covid Healthy Controls (PCHC) were recruited during the same time. Serum proteome of participants was determined 5 to 19 months following acute COVID-19. Like in the previous two cohorts, a factorial design was applied to address an, in this case albeit slight, age and sex specific imbalance. Proteome analysis was performed with a more conventional proteomic platform, using nano-flowrate, 30-min chromatographic gradient elution on an Ultimate 3000 RSLnano HPLC coupled to a Thermo Scientific Q-Exactive Plus mass spectrometer operating in data independent acquisition (DIA) mode (*17*), followed by raw-data processing using DIA-NN.

Statistical analysis of the Charité post mild acute COVID19 cohort (PACS (with ME/CFS and without) vs.– Post Covid Healthy Controls (PCHC)) revealed 27 differentially abundant proteins (at significance level alpha = 0.05 (fdr <= 0.33) and the estimated fold-change threshold (1.1, Figure 1F), of which 7 (CA1, SAA1, APOB, ITIH3, IGHV1-2, LRG1, CFI) are elevated. In this cohort, the only complement protein significantly upregulated, CFI, was correlated with BMI (*r* ∼ 0.56). Most of the 27 proteins that are significantly regulated by PACS were previously identified in COVID-19. Increased levels of SAA1 (Serum Amyloid A1), ITIH3 (Inter-Alpha-Trypsin Inhibitor Heavy Chain H3), CFI (Complement Factor I), LRG1 (Leucine-Rich Alpha-2-Glycoprotein 1) and decreased levels of Lumican (LUM), Fibronectin 1 (FN1), Multimerin-1 (MMRN1), Cartilage Oligomeric Matrix Protein (COMP), Transgelin 2 (TAGLN2), and Profilin 1 (PFN1) are consistent with protein regulation in severe acute patients (*11, 18, 19*). The proteins mirror the response in inflammation, acute phase, coagulation, extracellular matrix as well as activation and degradation of platelets. The proteome in these individuals could therefore indicate a persistent infection or persistent immune activation.

Thus, while our reanalysis of the Zurich cohort, the analysis of an analogous post severe acute COVID19 cohort, and a third cohort specifically focussing on PACS patients with fatigue and exertional intolerance subsequent to a mild acute phase, revealed interesting proteomic signatures, we can not substantiate a consistent activation of the complement system in PACS. Only two CS proteins (CFI and CFP) were found to be significantly upregulated in PACS patients, however only by one cohort each and with low effect size. To evaluate the role of age, sex, and BMI on the complement system in the different cohorts, we consequently compared the dependence of complement proteins on PACS, and the different demographic factors (Figure 1D). This illustration visualises that the dependence of BMI dominates over other demographic factors, and PACS, when it comes to the abundance of complement proteins in the plasma proteome. We must conclude that the strong signal which indicates a persistent activation of the complement system, is not a generalizable to in patients suffering from PACS, and that this signal may have emerged due to an imbalance in age and BMI between case and controls in an exploratory cohort.

## Data Availability

All data produced in the present work are contained in the manuscript.

## Acknowledgements

We thank the Pa-Covid19 study group at Charité Universitätsmedizin Berlin for contribution of data and biosamples for this study, as well as our colleagues for supporting data acquisition, and analysis. This work was supported by the Ministry of Education and Research (BMBF), as part of the National Research Initiative ‘Mass Spectrometry in Systems Medicine’ (MSCoreSys), under grant agreement number 01EP2201 (to MR) and 16LW0239K (to MM), under Grant 01EP2201 (to CS) as well as the. Weidenhammer-Zoebele Foundation, and the Berlin University Alliance (BUA Link Lab, 501_Massenspektrometrie, 501_Linklab).

## Methods

### Cohorts

#### Charité post severe acute COVID19

The Charité post severe acute COVID19 cohort is a subcohort of the Pa-COVID-19 study, a prospective observational study registered with German clinical trials registry (DRKS 00021688) aiming to provide a platform for clinical characterisation of acute and post-acute COVID-19 (*26*). The study was approved by the ethics committee of Charité - Universitätsmedizin Berlin (EA2/066/20) and conducted in accordance with the declaration of Helsinki. This analysis includes patients from the Pa-COVID-19 study, who were followed-up as outpatients after acute SARS-CoV-2 infection. A total of 130 patients with plasma samples collected at month 6 post SARS-CoV-2 infection were included in this analysis. Of these 104 were hospitalised during acute infection and 44 were treated at ICU. 26 patients were not hospitalised and treated as outpatients with mild SARS-CoV-2 infection. Symptom assessment at outpatient presentation was recorded by a physician as described previously (*10*). Individuals having at least two symptoms out of cough, dyspnea, fatigue, headache, chest pain, gastrointestinal or depression symptoms were considered to be in the PACS group. Gastrointestinal symptoms were assigned to individuals suffering at least from two symptoms out of diarrhoea, vomitus, stomachache and nausea, while depression symptoms were assigned if at least two symptoms out of fear, panic and major depression were assigned to the individual.

We assessed the effect of the covariates sex, age, BMI and severity during the acute phase (by WHO grade) and identified none of them to be associated with PACS: point-biserial correlation(age) = 0, Cramér’s V(sex) = 0.12, point-biserial correlation(BMI) = 0.02, Cramér’s V(WHO severity) = 0.16 and p-value(age, ANOVA) = 1.00, p-value(sex, Chi^2^ independency test) = 0.17, p-value(BMI, ANOVA) = 0.82, p-value(WHO Severity, Chi^2^ independency test) = 0.68.

#### Charité post mild acute COVID19

A total of 53 PACS patients from the Pa-COVID-19 study with persistent moderate to severe fatigue, exertional intolerance and PEM following mild to moderate COVID-19 were included in this study. Of these n=25 fulfilled diagnostic criteria of ME/CFS based on the Canadian Consensus Criteria (CCC) referred to as PCS/ME/CFS as described previously (*27*). Patients were excluded from this study in case of relevant comorbidities (*28*), evidence of organ dysfunction, or pre-existing fatigue. The demographic characteristics of the study cohorts are presented in detail in Supplementary Table 3. Serum samples were collected a median of eight months following SARS-CoV-2 infection. As control, 27 healthy non-PACS were recruited a median of seven months after mild to moderate COVID-19 during the same time span.

To compensate for the difference between ages and achieve equal female/male contributions we opted for factorial design and split patients into sex (female/male) – age (above / below 42 years) subgroups. The average age of 42 years was taken as a threshold for age binarization. Two PACS non-ME/CFS samples and one healthy non-PACS sample were excluded due to quality reasons. Final size of the cohort was 77 individuals.

### Sample preparation

Semi-automated in-solution digestion was performed as previously described for high throughput clinical proteomics (*11*). To reduce variability all stocks and stock plates were prepared in advance and stored at -80°C until use. Briefly, 5 μl of thawed samples were transferred to the denaturation and reduction solution (50 μl 8M Urea, 100 mM ammonium bicarbonate (ABC), 5 μl 50 mM dithiothreitol per well) mixed and incubated at 30°C for 60 minutes. Then 5 μl were transferred from the iodoacetamide stock solution plate (100 mM) to the sample plate and incubated in the dark at RT for 30 minutes before dilution with 100 mM ABC buffer (340 μl). 220 μl of this solution was transferred to the pre-made trypsin stock solution plate (12.5 μl, 0.1 μg/μl) and incubated at 37°C for 17 h (Benchmark Scientific Incu-Mixer MP4). For quenching formic acid (10% v/v, 25 μl) was added and for cleaning C18 solid phase extraction in 96-well plates (BioPureSPE Macro 96-Well, 100 mg PROTO C18, The Nest Group) was used. After drying under vacuum the eluent was reconstituted in 60 μl 0.1% formic acid. Before sample transferring to a new plate, insoluble particles were removed by centrifugation.

### Mass spectrometry and Computational proteomics

#### Charité post severe acute COVID19

5 μg plasma peptides were chromatographically separated in a 3-minute water to acetonitrile active gradient on an Agilent Infinity II HPLC coupled to a Sciex Triple TOF 6600 mass spectrometer operating in ScanningSWATH mode with minor changes in the liquid chromatography method (*13*).

Raw proteomics data was processed using DIA-NN, version 1.8 (*29*). The MS1 and MS2 mass accuracies were set to 20 ppm, and the scan window to 6. Peptide ions were annotated with a publicly available spectral library for human plasma (*30*) and spectra and RT information were replaced with DIA-NN deep learning-based prediction. Protein inference was switched off and the match-between-runs (MBR) option was enabled.

#### Charité post mild acute COVID19

Peptide separation was accomplished in a 35-minute water to acetonitrile gradient (buffer A: 0.1 % formic acid, buffer B: 80 % ACN, 0.1 % formic acid) on an Ultimate 3000 RSLnanoHPLC coupled to a Q-Exactive Plus mass spectrometer (both ThermoFisher Scientific) operating in data independent acquisition (DIA) mode (*17*).

1 μg peptide mixtures were analysed by a two-linear-column system. Separation was done on a C18 column (Acclaim PepMap C18, 2 μm; 100 Å; 75μm, 15cm length, Thermo Fisher Scientific) with an active gradient from 4-38% buffer B in 30 min. To generate a spectral library, a pool of study samples was fractionated using a high pH fractionation kit according to manufacturer instructions (Pierce 84868). Fractionated samples were concentrated and separated as described above in a longer linear gradient from 5-28% buffer B in 63 min. Total acquisition time was 100 min.

The data was annotated with DIA-NN 1.8.1 (*29*) to the human reference proteome (Uniprot UP000005640_9606, accessed 2023-01-17) in library free mode using standard settings. Raw data were processed using DIA-NN 1.8.1 with scan window size set to 7 and MS2 and MS1 mass accuracies set to 20 and 10 ppm, respectively. Additionally, the match between-runs (MBR) option was enabled. The output was filtered at 1% FDR on peptide level.

### Data processing

#### Charité post severe acute COVID19

Only proteotypic precursors with a Q-Value, Global Q-Value and Library Q-Value below 0.01 were used for further analysis. Precursors, which were only identified but could not be quantified due to low abundance, as well as precursors that were present in less than a third of all samples containing a precursor of the same protein were also excluded from analysis. The sample precursor quantity distributions were normalized to the median precursor abundance across all samples using precursors present in at least 90% of all samples. Missing values were imputed iteratively first across samples during the same visite from individuals exhibiting the same severity (WHO grade) during acute phase for precursors missing in up to one third of the samples per group, then over the complete 6 month followup visit for samples missing in up to one third of the samples per visit and last over all visits from acute phase till 12 months follow-up using KNNImputer with k=5 from the scikit-learn impute package for Python (*31*). If no precursor of a given protein was identified in the raw data of a sample, then they were only imputed on protein level after summarization of precursor to protein quantities. The resulting precursors x samples matrix was filtered on outlier samples by applying quality control filters on the number of identified precursors, precursor intensity distributions and technical parameter collected in the DIA-NN stats-report. After exclusion of samples with insufficient quality, the filtered raw precursor quantities were again normalised, imputed as described above and summarised to protein quantities using the MaxLFQ algorithm (*32*) implemented by DIA-NN (*29, 33*). Finally, missing values on protein level were imputed with KNNImputer with k=5 from the scikit-learn impute package for Python (*3*1). The final protein matrix contained 200 proteins and 120 samples (Supplementary Table 4).

#### Charité post mild acute COVID19

For data integration we used DIA-NN (Demichev et al, 2020) output matrix of normalised precursor intensities for 80 study samples. After quality control three samples were identified as outliers and excluded, thus leaving for preprocessing 77 samples. Following pre-processing steps were applied: Cyclic loess group-wise (group levels: PCS(26 samples), PCS/ME/CFS (25 samples), PCHC (26 samples)) pre-normalization (*34*), implemented in the R package LIMMA (*35*) with option “fast” (*36*), stepwise imputation of missing values in peptides having completeness over 66%, using bpca method (*37*) implemented in R package pcaMethods (*38*), total set based cyclic loess normalisation (*34*) and PLM summarisation (*39*) implemented in R package preprocessCore (*40*) to get protein level data. Final protein matrix size was 233 proteins, 77 samples (Supplementary Table 5).

#### Statistical Analysis

Statistical analysis of proteomics data was carried out in R using publicly available packages. Balancing was achieved per cohort by splitting all patients into disease subgroups by sex – female or male and by age – below or above the mean age of individuals. For the Charité post severe acute COVID19 cohort the threshold for age subgroups was set to 52 instead of the mean of 58 for better comparison with the other two datasets. We ensured a sufficient number of individuals per subgroup. Individuals with too different age and/or BMI were excluded per dataset. In this way it was possible to attain reasonable balancing of age and BMI in disease age-sex subgroups. The final balanced Zurich cohort consisted of 85 individuals (56 non-PACS + 29 PACS with approximately the same odds ratio PACS to non-PACS as in the initial data set), the Charité post severe acute COVID19 cohort of 120 individuals (61 PACS to 59 non-PACS) and the Charité post mild acute COVID19 cohort of 77 individuals (25 PACS/ME/CFS, 26 PACS/non-ME/CFS and 26 non-PACS). The demographic characteristics for the three balanced cohorts are summarised in Supplementary Tables 1-3.

For analysis of all datasets, we used moderated statistics implemented in R package LIMMA (Ritchie, 2015) – due to borrowing information across features it is stable even for experiments with small numbers of samples in a group. We applied the same model to the unbalanced Zurich cohort as described by the authors (log2(*P*) is the log2-transformed expression of a protein: log2(P) ∼ 0 + Diagnosis + Age + Sex + Severity). To the balanced cohort as well as to the two validation cohorts we applied a different model accounting age and sex effects and including an additional BMI term: log2(*P*) ∼ *0 + Diagnosis _Sex_Age + BMI + Severity*. Factor *Severity* was not included into the ME/CFS cohort model, as the samples were of mild to moderate severity. In the balanced model the PACS – non-PACS difference was calculated as equally weighted average of subgroup differences (below F/M stands for female/male, AMA/BMA for age above/below mean age):

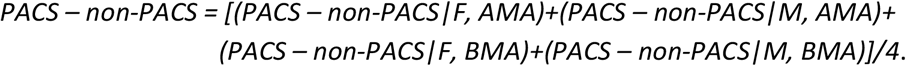

In the Charité post mild acute COVID19 cohort the categorical factor Diagnosis_Sex_Age had three levels (see Supplementary Table 3): PCS, PCS/ME/CFS, and PCHC; and was further split into Sex – Age subgroups. In this model the PACS – non-PACS difference was calculated as equally weighted average of eight subgroup differences (below ME stands for ME/CFS, F/M stands for female/male, A/B42 for age above/below 42):

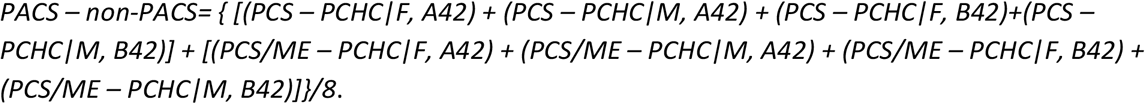

For finding regulated features we applied the following criteria: significance level alpha was set to 0.05, which was sufficient to guarantee fdr < 0.33 in contrast *PACS – non-PACS*. The fold change threshold FCT=1.1 was set to guarantee that the measured signal is above the noise level. As such we have taken half of the median residual standard deviation of the linear model. It was also higher than the coefficient of variation in quality control samples. To make cross datasets comparisons easier, we applied the same thresholds to all three datasets.

## References

1. Global Burden of Disease Long COVID Collaboratorss, S. Wulf Hanson, C. Abbafati, J. G. Aerts, Z. Al-Aly, C. Ashbaugh, T. Ballouz, O. Blyuss, P. Bobkova, G. Bonsel, S. Borzakova, D. Buonsenso, D. Butnaru, A. Carter, H. Chu, C. De Rose, M. M. Diab, E. Ekbom, M. El Tantawi, V. Fomin, R. Frithiof, A. Gamirova, P. V. Glybochko, J. A. Haagsma, S. Haghjooy Javanmard, E. B. Hamilton, G. Harris, M. H. Heijenbrok-Kal, R. Helbok, M. E. Hellemons, D. Hillus, S. M. Huijts, M. Hultström, W. Jassat, F. Kurth, I.-M. Larsson, M. Lipcsey, C. Liu, C. D. Loflin, A. Malinovschi, W. Mao, L. Mazankova, D. McCulloch, D. Menges, N. Mohammadifard, D. Munblit, N. A. Nekliudov, O. Ogbuoji, I. M. Osmanov, J. L. Peñalvo, M. S. Petersen, M. A. Puhan, M. Rahman, V. Rass, N. Reinig, G. M. Ribbers, A. Ricchiuto, S. Rubertsson, E. Samitova, N. Sarrafzadegan, A. Shikhaleva, K. E. Simpson, D. Sinatti, J. B. Soriano, E. Spiridonova, F. Steinbeis, A. A. Svistunov, P. Valentini, B. J. van de Water, R. van den Berg-Emons, E. Wallin, M. Witzenrath, Y. Wu, H. Xu, T. Zoller, C. Adolph, J. Albright, J. O. Amlag, A. Y. Aravkin, B. L. Bang-Jensen, C. Bisignano, R. Castellano, E. Castro, S. Chakrabarti, J. K. Collins, X. Dai, F. Daoud, C. Dapper, A. Deen, B. B. Duncan, M. Erickson, S. B. Ewald, A. J. Ferrari, A. D. Flaxman, N. Fullman, A. Gamkrelidze, J. R. Giles, G. Guo, S. I. Hay, J. He, M. Helak, E. N. Hulland, M. Kereselidze, K. J. Krohn, A. Lazzar-Atwood, A. Lindstrom, R. Lozano, D. C. Malta, J. Månsson, A. M. Mantilla Herrera, A. H. Mokdad, L. Monasta, S. Nomura, M. Pasovic, D. M. Pigott, R. C. Reiner Jr, G. Reinke, A. L. P. Ribeiro, D. F. Santomauro, A. Sholokhov, E. E. Spurlock, R. Walcott, A. Walker, C. S. Wiysonge, P. Zheng, J. P. Bettger, C. J. L. Murray, T. Vos, Estimated global proportions of individuals with persistent fatigue, cognitive, and respiratory symptom clusters following symptomatic COVID-19 in 2020 and 2021. JAMA 328, 1604–1615 (2022).

2. H. Brüssow, K. Timmis, COVID-19: long covid and its societal consequences. Environ. Microbiol. 23, 4077–4091 (2021).

3. A. Gandjour, Long COVID: Costs for the German economy and health care and pension system. BMC Health Serv. Res. 23, 641 (2023).

4. H. E. Davis, L. McCorkell, J. M. Vogel, E. J. Topol, Long COVID: major findings, mechanisms and recommendations. Nat. Rev. Microbiol. 21, 133–146 (2023).

5. C. Cervia-Hasler, S. C. Brüningk, T. Hoch, B. Fan, G. Muzio, R. C. Thompson, L. Ceglarek, R. Meledin, P. Westermann, M. Emmenegger, P. Taeschler, Y. Zurbuchen, M. Pons, D. Menges, T. Ballouz, S. Cervia-Hasler, S. Adamo, M. Merad, A. W. Charney, M. Puhan, P. Brodin, J. Nilsson, A. Aguzzi, M. E. Raeber, C. B. Messner, N. D. Beckmann, K. Borgwardt, O. Boyman, Persistent complement dysregulation with signs of thromboinflammation in active Long Covid. Science 383, eadg7942 (2024).

6. V. Tsampasian, H. Elghazaly, R. Chattopadhyay, M. Debski, T. K. P. Naing, P. Garg, A. Clark, E. Ntatsaki, V. S. Vassiliou, Risk Factors Associated With Post-COVID-19 Condition: A Systematic Review and Meta-analysis. JAMA Intern. Med. 183, 566–580 (2023).

7. M. Gaya da Costa, F. Poppelaars, C. van Kooten, T. E. Mollnes, F. Tedesco, R. Würzner, L. A. Trouw, L. Truedsson, M. R. Daha, A. Roos, M. A. Seelen, Age and Sex-Associated Changes of Complement Activity and Complement Levels in a Healthy Caucasian Population. Front. Immunol. 9, 2664 (2018).

8. N. Dordevic, C. Dierks, E. Hantikainen, V. Farztdinov, F. Amari, V. Verri Hernandes, A. De Grandi, F. S. Domingues, M. Mülleder, P. P. Pramstaller, J. Rainer, M. Ralser, Pervasive influence of hormonal contraceptives on the human plasma proteome in a broad population study, bioRxiv (2023). 10.1101/2023.10.11.23296871.

9. P. Durrington, Blood lipids after COVID-19 infection, The lancet. Diabetes & endocrinology. 11 (2023)pp. 68–69.

10. F. Steinbeis, C. Thibeault, F. Doellinger, R. M. Ring, M. Mittermaier, C. Ruwwe-Glösenkamp, F. Alius, P. Knape, H.-J. Meyer, L. J. Lippert, E. T. Helbig, D. Grund, B. Temmesfeld-Wollbrück, N. Suttorp, L. E. Sander, F. Kurth, T. Penzkofer, M. Witzenrath, T. Zoller, Severity of respiratory failure and computed chest tomography in acute COVID-19 correlates with pulmonary function and respiratory symptoms after infection with SARS-CoV-2: An observational longitudinal study over 12 months. Respir. Med. 191, 106709 (2022).

11. C. B. Messner, V. Demichev, D. Wendisch, L. Michalick, M. White, A. Freiwald, K. Textoris-Taube, S. I. Vernardis, A.-S. Egger, M. Kreidl, D. Ludwig, C. Kilian, F. Agostini, A. Zelezniak, C. Thibeault, M. Pfeiffer, S. Hippenstiel, A. Hocke, C. von Kalle, A. Campbell, C. Hayward, D. J. Porteous, R. E. Marioni, C. Langenberg, K. S. Lilley, W. M. Kuebler, M. Mülleder, C. Drosten, M. Witzenrath, F. Kurth, L. E. Sander, M. Ralser, Ultra-high-throughput clinical proteomics reveals classifiers of COVID-19 infection. Cell Systems, doi:10.1016/j.cels.2020.05.012 (2020).

12. C. B. Messner, V. Demichev, J. Muenzner, S. K. Aulakh, N. Barthel, A. Röhl, L. Herrera-Domínguez, A.-S. Egger, S. Kamrad, J. Hou, G. Tan, O. Lemke, E. Calvani, L. Szyrwiel, M. Mülleder, K. S. Lilley, C. Boone, G. Kustatscher, M. Ralser, The proteomic landscape of genome-wide genetic perturbations. Cell 186, 2018–2034.e21 (2023).

13. C. B. Messner, V. Demichev, N. Bloomfield, J. S. L. Yu, M. White, M. Kreidl, A.-S. Egger, A. Freiwald, G. Ivosev, F. Wasim, A. Zelezniak, L. Jürgens, N. Suttorp, L. E. Sander, F. Kurth, K. S. Lilley, M. Mülleder, S. Tate, M. Ralser, Ultra-fast proteomics with Scanning SWATH. Nat. Biotechnol. 39, 846–854 (2021).

14. K. Kostka, E. Roel, N. T. H. Trinh, N. Mercadé-Besora, A. Delmestri, L. Mateu, R. Paredes, T. Duarte-Salles, D. Prieto-Alhambra, M. Català, A. M. Jödicke, “The burden of post-acute COVID-19 symptoms in a multinational network cohort analysis.” Nat. Commun. 14, 7449 (2023).

15. F. Legler, L. Meyer-Arndt, L. Mödl, C. Kedor, H. Freitag, E. Stein, U. Hoppmann, R. Rust, K. Wittke, N. Siebert, J. Behrens, A. Thiel, F. Konietschke, F. Paul, C. Scheibenbogen, J. Bellmann-Strobl, Long-term symptom severity and clinical biomarkers in post-COVID-19/chronic fatigue syndrome: results from a prospective observational cohort. EClinicalMedicine 63, 102146 (2023).

16. Committee on the Diagnostic Criteria for Myalgic Encephalomyelitis/Chronic Fatigue Syndrome, Board on the Health of Select Populations, Institute of Medicine, Current Case Definitions and Diagnostic Criteria, Terminology, and Symptom Constructs and Clusters (National Academies Press (US), 2015).

17. J. Doellinger, C. Blumenscheit, A. Schneider, P. Lasch, Increasing Proteome Depth While Maintaining Quantitative Precision in Short-Gradient Data-Independent Acquisition Proteomics. J. Proteome Res. 22, 2131–2140 (2023).

18. V. Demichev, P. Tober-Lau, O. Lemke, T. Nazarenko, C. Thibeault, H. Whitwell, A. Röhl, A. Freiwald, L. Szyrwiel, D. Ludwig, C. Correia-Melo, S. K. Aulakh, E. T. Helbig, P. Stubbemann, L. J. Lippert, N.-M. Grüning, O. Blyuss, S. Vernardis, M. White, C. B. Messner, M. Joannidis, T. Sonnweber, S. J. Klein, A. Pizzini, Y. Wohlfarter, S. Sahanic, R. Hilbe, B. Schaefer, S. Wagner, M. Mittermaier, F. Machleidt, C. Garcia, C. Ruwwe-Glösenkamp, T. Lingscheid, L. Bosquillon de Jarcy, M. S. Stegemann, M. Pfeiffer, L. Jürgens, S. Denker, D. Zickler, P. Enghard, A. Zelezniak, A. Campbell, C. Hayward, D. J. Porteous, R. E. Marioni, A. Uhrig, H. Müller-Redetzky, H. Zoller, J. Löffler-Ragg, M. A. Keller, I. Tancevski, J. F. Timms, A. Zaikin, S. Hippenstiel, M. Ramharter, M. Witzenrath, N. Suttorp, K. Lilley, M. Mülleder, L. E. Sander, M. Ralser, F. Kurth, M. Kleinschmidt, K. M. Heim, B. Millet, L. Meyer-Arndt, R. H. Hübner, T. Andermann, J. M. Doehn, B. Opitz, B. Sawitzki, D. Grund, P. Radünzel, M. Schürmann, T. Zoller, F. Alius, P. Knape, A. Breitbart, Y. Li, F. Bremer, P. Pergantis, D. Schürmann, B. Temmesfeld-Wollbrück, D. Wendisch, S. Brumhard, S. S. Haenel, C. Conrad, P. Georg, K.-U. Eckardt, L. Lehner, J. M. Kruse, C. Ferse, R. Körner, C. Spies, A. Edel, S. Weber-Carstens, A. Krannich, S. Zvorc, L. Li, U. Behrens, S. Schmidt, M. Rönnefarth, C. Dang-Heine, R. Röhle, E. Lieker, L. Kretzler, I. Wirsching, C. Wollboldt, Y. Wu, G. Schwanitz, D. Hillus, S. Kasper, N. Olk, A. Horn, D. Briesemeister, D. Treue, M. Hummel, V. M. Corman, C. Drosten, C. von Kalle, A time-resolved proteomic and prognostic map of COVID-19. Cell Syst. 12, 780–794.e7 (2021).

19. B. Shen, X. Yi, Y. Sun, X. Bi, J. Du, C. Zhang, S. Quan, F. Zhang, R. Sun, L. Qian, W. Ge, W. Liu, S. Liang, H. Chen, Y. Zhang, J. Li, J. Xu, Z. He, B. Chen, J. Wang, H. Yan, Y. Zheng, D. Wang, J. Zhu, Z. Kong, Z. Kang, X. Liang, X. Ding, G. Ruan, N. Xiang, X. Cai, H. Gao, L. Li, S. Li, Q. Xiao, T. Lu, Y. Zhu, H. Liu, H. Chen, T. Guo, Proteomic and Metabolomic Characterization of COVID-19 Patient Sera. Cell 182, 59–72.e15 (2020).

20. M. Yuan, D. Huang, C.-C. D. Lee, N. C. Wu, A. M. Jackson, X. Zhu, H. Liu, L. Peng, M. J. van Gils, R. W. Sanders, D. R. Burton, S. M. Reincke, H. Prüss, J. Kreye, D. Nemazee, A. B. Ward, I. A. Wilson, Structural and functional ramifications of antigenic drift in recent SARS-CoV-2 variants. Science 373, 818–823 (2021).

21. F. Duan, Y. Wang, T. Chen, Z. Zhu, M. Yu, H. Dai, S. Zheng, Y. Lu, T. Li, X. Qiu, A novel strategy for identifying biomarker in serum of patient with COVID-19 using immune complex. Signal Transduct Target Ther 7, 63 (2022).

22. S. Pushalkar, S. Wu, S. Maity, M. Pressler, J. Rendleman, B. Vitrinel, L. Jeffery, R. Abdelhadi, M. Chen, T. Ross, M. Carlock, H. Choi, C. Vogel, Complex changes in serum protein levels in COVID-19 convalescents. Sci. Rep. 14, 4479 (2024).

23. R. Martins-Gonçalves, E. D. Hottz, P. T. Bozza, Acute to post-acute COVID-19 thromboinflammation persistence: Mechanisms and potential consequences. Curr Res Immunol 4, 100058 (2023).

24. M. Ashburner, C. A. Ball, J. A. Blake, D. Botstein, H. Butler, J. M. Cherry, A. P. Davis, K. Dolinski, S. S. Dwight, J. T. Eppig, M. A. Harris, D. P. Hill, L. Issel-Tarver, A. Kasarskis, S. Lewis, J. C. Matese, J. E. Richardson, M. Ringwald, G. M. Rubin, G. Sherlock, Gene ontology: tool for the unification of biology. The Gene Ontology Consortium. Nat. Genet. 25, 25–29 (2000).

25. Gene Ontology Consortium, S. A. Aleksander, J. Balhoff, S. Carbon, J. M. Cherry, H. J. Drabkin, D. Ebert, M. Feuermann, P. Gaudet, N. L. Harris, D. P. Hill, R. Lee, H. Mi, S. Moxon, C. J. Mungall, A. Muruganugan, T. Mushayahama, P. W. Sternberg, P. D. Thomas, K. Van Auken, J. Ramsey, D. A. Siegele, R. L. Chisholm, P. Fey, M. C. Aspromonte, M. V. Nugnes, F. Quaglia, S. Tosatto, M. Giglio, S. Nadendla, G. Antonazzo, H. Attrill, G. Dos Santos, S. Marygold, V. Strelets, C. J. Tabone, J. Thurmond, P. Zhou, S. H. Ahmed, P. Asanitthong, D. Luna Buitrago, M. N. Erdol, M. C. Gage, M. Ali Kadhum, K. Y. C. Li, M. Long, A. Michalak, A. Pesala, A. Pritazahra, S. C. C. Saverimuttu, R. Su, K. E. Thurlow, R. C. Lovering, C. Logie, S. Oliferenko, J. Blake, K. Christie, L. Corbani, M. E. Dolan, H. J. Drabkin, D. P. Hill, L. Ni, D. Sitnikov, C. Smith, A. Cuzick, J. Seager, L. Cooper, J. Elser, P. Jaiswal, P. Gupta, P. Jaiswal, S. Naithani, M. Lera-Ramirez, K. Rutherford, V. Wood, J. L. De Pons, M. R. Dwinell, G. T. Hayman, M. L. Kaldunski, A. E. Kwitek, S. J. F. Laulederkind, M. A. Tutaj, M. Vedi, S.-J. Wang, P. D’Eustachio, L. Aimo, K. Axelsen, A. Bridge, N. Hyka-Nouspikel, A. Morgat, S. A. Aleksander, J. M. Cherry, S. R. Engel, K. Karra, S. R. Miyasato, R. S. Nash, M. S. Skrzypek, S. Weng, E. D. Wong, E. Bakker, T. Z. Berardini, L. Reiser, A. Auchincloss, K. Axelsen, G. Argoud-Puy, M.-C. Blatter, E. Boutet, L. Breuza, A. Bridge, C. Casals-Casas, E. Coudert, A. Estreicher, M. Livia Famiglietti, M. Feuermann, A. Gos, N. Gruaz-Gumowski, C. Hulo, N. Hyka-Nouspikel, F. Jungo, P. Le Mercier, D. Lieberherr, P. Masson, A. Morgat, I. Pedruzzi, L. Pourcel, S. Poux, C. Rivoire, S. Sundaram, A. Bateman, E. Bowler-Barnett, H. Bye-A-Jee, P. Denny, A. Ignatchenko, R. Ishtiaq, A. Lock, Y. Lussi, M. Magrane, M. J. Martin, S. Orchard, P. Raposo, E. Speretta, N. Tyagi, K. Warner, R. Zaru, A. D. Diehl, R. Lee, J. Chan, S. Diamantakis, D. Raciti, M. Zarowiecki, M. Fisher, C. James-Zorn, V. Ponferrada, A. Zorn, S. Ramachandran, L. Ruzicka, M. Westerfield, The Gene Ontology knowledgebase in 2023. Genetics 224 (2023).

26. F. Kurth, M. Roennefarth, C. Thibeault, V. M. Corman, H. Müller-Redetzky, M. Mittermaier, C. Ruwwe-Glösenkamp, K. M. Heim, A. Krannich, S. Zvorc, S. Schmidt, L. Kretzler, C. Dang-Heine, M. Rose, M. Hummel, A. Hocke, R. H. Hübner, B. Opitz, M. A. Mall, J. Röhmel, U. Landmesser, B. Pieske, S. Knauss, M. Endres, J. Spranger, F. P. Mockenhaupt, F. Tacke, S. Treskatsch, S. Angermair, B. Siegmund, C. Spies, S. Weber-Carstens, K.-U. Eckardt, D. Schürmann, A. Uhrig, M. S. Stegemann, T. Zoller, C. Drosten, N. Suttorp, M. Witzenrath, S. Hippenstiel, C. von Kalle, L. E. Sander, Studying the pathophysiology of coronavirus disease 2019: a protocol for the Berlin prospective COVID-19 patient cohort (Pa-COVID-19). Infection 48, 619–626 (2020).

27. C. Kedor, H. Freitag, L. Meyer-Arndt, K. Wittke, L. G. Hanitsch, T. Zoller, F. Steinbeis, M. Haffke, G. Rudolf, B. Heidecker, T. Bobbert, J. Spranger, H.-D. Volk, C. Skurk, F. Konietschke, F. Paul, U. Behrends, J. Bellmann-Strobl, C. Scheibenbogen, A prospective observational study of post-COVID-19 chronic fatigue syndrome following the first pandemic wave in Germany and biomarkers associated with symptom severity. Nat. Commun. 13, 5104 (2022).

28. L. Nacul, F. J. Authier, C. Scheibenbogen, L. Lorusso, I. B. Helland, J. A. Martin, C. A. Sirbu, A. M. Mengshoel, O. Polo, U. Behrends, H. Nielsen, P. Grabowski, S. Sekulic, N. Sepulveda, F. Estévez-López, P. Zalewski, D. F. H. Pheby, J. Castro-Marrero, G. K. Sakkas, E. Capelli, I. Brundsdlund, J. Cullinan, A. Krumina, J. Bergquist, M. Murovska, R. C. W. Vermuelen, E. M. Lacerda, European Network on Myalgic Encephalomyelitis/Chronic Fatigue Syndrome (EUROMENE): Expert Consensus on the Diagnosis, Service Provision, and Care of People with ME/CFS in Europe. Medicina 57 (2021).

29. V. Demichev, C. B. Messner, S. I. Vernardis, K. S. Lilley, M. Ralser, DIA-NN: neural networks and interference correction enable deep proteome coverage in high throughput. Nat. Methods 17, 41–44 (2020).

30. R. Bruderer, J. Muntel, S. Müller, O. M. Bernhardt, T. Gandhi, O. Cominetti, C. Macron, J. Carayol, O. Rinner, A. Astrup, W. H. M. Saris, J. Hager, A. Valsesia, L. Dayon, L. Reiter, Analysis of 1508 Plasma Samples by Capillary-Flow Data-Independent Acquisition Profiles Proteomics of Weight Loss and Maintenance. Mol. Cell. Proteomics 18, 1242–1254 (2019).

31. F. Pedregosa, G. Varoquaux, A. Gramfort, V. Michel, B. Thirion, O. Grisel, M. Blondel, P. Prettenhofer, R. Weiss, V. Dubourg, Others, Scikit-learn: Machine learning in Python. the Journal of machine Learning research 12, 2825–2830 (2011).

32. J. Cox, M. Y. Hein, C. A. Luber, I. Paron, N. Nagaraj, M. Mann, Accurate proteome-wide label-free quantification by delayed normalization and maximal peptide ratio extraction, termed MaxLFQ. Mol. Cell. Proteomics 13, 2513–2526 (2014).

33. F. Kistner, J. L. Grossmann, L. R. Sinn, V. Demichev, QuantUMS: uncertainty minimisation enables confident quantification in proteomics, bioRxiv (2023)p. 2023.06.20.545604.

34. B. M. Bolstad, R. A. Irizarry, M. Astrand, T. P. Speed, A comparison of normalization methods for high density oligonucleotide array data based on variance and bias. Bioinformatics 19, 185–193 (2003).

35. M. E. Ritchie, B. Phipson, D. Wu, Y. Hu, C. W. Law, W. Shi, G. K. Smyth, limma powers differential expression analyses for RNA-sequencing and microarray studies. Nucleic Acids Res. 43, e47 (2015).

36. K. V. Ballman, D. E. Grill, A. L. Oberg, T. M. Therneau, Faster cyclic loess: normalizing RNA arrays via linear models. Bioinformatics 20, 2778–2786 (2004).

37. S. Oba, M.-A. Sato, I. Takemasa, M. Monden, K.-I. Matsubara, S. Ishii, A Bayesian missing value estimation method for gene expression profile data. Bioinformatics 19, 2088–2096 (2003).

38. W. Stacklies, H. Redestig, M. Scholz, D. Walther, J. Selbig, pcaMethods--a bioconductor package providing PCA methods for incomplete data. Bioinformatics 23, 1164–1167 (2007).

39. B. Bolstad, “Preprocessing and Normalization for Affymetrix GeneChip Expression Microarrays” in Methods in Microarray Normalization, P. Stafford, Ed. (Taylor & Francis, Boca Raton, FL, 2008), pp. 41–59.

40. B. Bolstad, preprocessCore: A collection of pre-processing functions (2021). https://rdrr.io/bioc/preprocessCore/.

